# Aperiodic neural activity is a biomarker for depression severity

**DOI:** 10.1101/2023.11.07.23298040

**Authors:** C. Hacker, M.M Mocchi, J. Xiao, B.A. Metzger, J.A. Adkinson, B.R. Pascuzzi, R.C. Mathura, D. Oswalt, A. Watrous, E. Bartoli, A. Allawala, V. Pirtle, X. Fan, I. Danstrom, B. Shofty, G. Banks, Y. Zhang, M. Armenta-Salas, K. Mirpour, N. Provenza, S. Mathew, J. Cohn, D. Borton, W. Goodman, N. Pouratian, S.A. Sheth, K.R. Bijanki

**Affiliations:** Baylor College of Medicine Department of Neurosurgery; Washington University in St. Louis Department of Neurosurgery; University of Pennsylvania Department of Neurosurgery; Brown University Department of Biomedical Engineering and Carney Institute for Brain Science; Brown University Department of Veterans Affairs Center for Neurorestoration and Neurotechnology; University of Texas Southwestern, Department of Neurosurgery; Baylor College of Medicine Department of Psychiatry; University of Pittsburgh Department of Psychology

**Keywords:** aperiodic slope exponent mood measure quantification affective vmPFC amygdala treatment-resistant depression

## Abstract

A reliable physiological biomarker for Major Depressive Disorder (MDD) is necessary to improve treatment success rates by shoring up variability in outcome measures. In this study, we establish a passive biomarker that tracks with changes in mood on the order of minutes to hours. We record from intracranial electrodes implanted deep in the brain – a surgical setting providing exquisite temporal and spatial sensitivity to detect this relationship in a difficult-to-measure brain area, the ventromedial prefrontal cortex (VMPFC). The aperiodic slope of the power spectral density captures the balance of activity across all frequency bands and is construed as a putative proxy for excitatory/inhibitory balance in the brain. This study demonstrates how shifts in aperiodic slope correlate with depression severity in a clinical trial of deep brain stimulation for treatment-resistant depression (TRD). The correlation between depression severity scores and aperiodic slope is significant in N=5 subjects, indicating that flatter (less negative) slopes correspond to reduced depression severity, especially in the ventromedial prefrontal cortex. This biomarker offers a new way to track patient response to MDD treatment, facilitating individualized therapies in both intracranial and non-invasive monitoring scenarios.

**One sentence summary:** The aperiodic component of the power spectral density robustly tracks depression severity on the order of minutes to hours.

---

Major Depressive Disorder (MDD) is a debilitating disorder with a poor treatment success rate despite its annual global cost approaching $92.7 billion^1^. Around one in three individuals experience no meaningful symptom improvement after multiple therapeutic attempts, and are diagnosed with treatment-resistant depression (TRD)^2^. To improve the therapeutic process, a reliable biomarker for depression could aid in selecting optimal therapeutic modalities, as well as enhance the tracking of symptom improvement and treatment efficacy. The current reliance on clinician-administered scales for monitoring changes in symptoms has limitations, including impaired expression of emotional experience (alexithymia), environmental influences, and socio-cultural factors^3^. Introducing a physiological biomarker would provide a more stable means of tracking symptoms, screening potential therapeutic approaches, and quantifying therapeutic efficacy.

The emergence of electrical stimulation-based treatments like transcranial magnetic stimulation (TMS) and deep brain stimulation (DBS) has offered hope to patients with unmet clinical needs and presents an opportunity for direct brain recording, potentially facilitating the identification of a physiological biomarker in neural electrophysiology. Previous efforts to find a biomarker have primarily explored fluctuations in oscillatory power bands within specific affective or reward processing regions during affective tasks^4–9^ to assess the efficacy of MDD and TRD treatment. However, oscillatory markers for DBS treatment tracking or mood diagnostics exhibit substantial inconsistency in frequency range and anatomical location across patients and studies, or may only be detected after weeks to months of treatment^7,9–11^. A biomarker that robustly detects changes in mood more immediately would allow for much quicker therapeutic titration or determination of non-efficacy which would allow patients to more rapidly progress through the therapeutic algorithm space to identify their own optimal treatment.

The aperiodic component (exponent) of power spectral density, known as the 1/f slope, is an underexplored biological signal with potential significance. Typically considered ‘noise’ and removed in data processing, recent research highlights its ability to quantify the excitatory to inhibitory (E/I) balance in the brain^12^. As a ‘snapshot’ of activity across all frequency bands, we hypothesize that aperiodic slope in the affective network may represent the degree of effective coordination of activity across the distributed brain regions involved in maintaining healthy affect. Variations in the aperiodic slope are linked to age differences^13^, diseases^14^, and depression severity between subjects^15^. Notably, a flattening of the aperiodic slope is observed in patients receiving pharmacological treatment for MDD^16^, though this study did not distinguish between responders and non-responders. Aperiodic slope was shown to change following weeks to months of subgenual cingulate DBS therapy^17^. While prior studies have demonstrated its capacity to capture long-term mood changes, the potential for tracking depressive symptom severity with high temporal precision within subjects remains unexplored. The aperiodic slope has shown rapid shifts in response to visual and auditory stimuli^18,19^, suggesting its potential for tracking mood changes at a faster pace than previously investigated.

In this study, our objective is to establish the aperiodic slope as a physiological biomarker for Major Depressive Disorder (MDD) on the order of minutes to hours. We investigate whether the aperiodic slope correlates with depression severity, as measured by a rapid adaptive depression inventory^20^, in patients participating in a clinical trial of deep brain stimulation (DBS) for treatment-resistant depression (TRD) (n = 5). Depression severity scores were collected multiple times per day throughout the 9-day in-patient stay, ranging from minutes to hours apart. A consistent physiological biomarker that exhibits high temporal resolution within and between patients would enable more efficient screening of multiple therapeutic approaches, facilitating the identification of optimal treatment strategies for each individual and ultimately moving the field toward the goal of achieving active control of the dysfunctional affective system in TRD.

## Methods

### Study design

Patient enrollment and depression assessment was equivalent to procedures described in Xiao et al., 2023. Briefly, TRD subjects (n=5) were enrolled in a clinical trial (NCT03437928) of individualized deep brain stimulation approved by the Baylor College of Medicine Institutional Review Board. Each subject was implanted with permanent deep brain stimulation leads and temporary stereoelectroencephalography (sEEG) recording electrodes in depression-relevant prefrontal regions. Following surgical implant, subjects underwent intensive inpatient monitoring for 9 days, during which frequent measures of depression severity were obtained using the Computerized Adaptive Test–Depression Inventory (CAT-DI)^20^. CAT-DI delivers a series of individual questions indexing the patient’s current mood state, adaptively sampling from a large question bank based on prior responses and computes a summary estimate of disease severity (0-100) with scores categorized as normal (<50), mild (50-65), moderate (66-75), or severe (>75). CAT-DI has high test-retest reliability (r = .92), strong correlation (r = 0.75 vs. HAM-D, r = 0.81 vs. PHQ-9) to established clinical scales^20,21^, and is typically completed within 1-3 minutes. CAT-DI typically asks patient mood over a time frame of 2 weeks. For the current study, the time frame was customized to reflect a 1-hour period.

### Intracranial recordings

Subsequent to each depression severity (CAT-DI) measurement, signals were recorded from sEEG electrodes while the patient fixated on a crosshair for approximately 1 to 3 minutes. Signals were recorded with a Cerebus data acquisition system (BlackRock Microsystems) at 2 kHz. Some recordings in patient five were recorded at 30kHz for reasons pertaining to other studies (in which case signals were resampled by decimation to 2 kHz after application of a lowpass Chebyshev Type I infinite impulse response (IIR) filter of order 8). All analyses were performed in MATLAB (Mathworks, inc.). Signals were inspected for artifact in the time and frequency domains. Temporal periods with excessive noise across channels were excluded. Individual electrodes with excessive line noise were removed from analyses. Two channels from intracranial leads were used as reference and ground and were thus not included in analysis. Remaining sEEG channels were re-referenced by subtracting the common median across channels at each time point. No stimulation was delivered during any of the recordings. Power spectral density estimation was performed with Welch’s method using one second windows with 50% overlap. Spectra were resampled to logarithmically spaced frequency bins and a linear fit was performed in log-log (log power and log frequency) over the 20-45 Hz range using the polyfit function with a degree of one^22^. The slope of this fit was used to compute correlations between spectral slope and depression severity. For retrospective biomarker optimization (**Figs. 2C and 3A**), the same fitting procedure was performed, parametric in bin edge frequencies.

### Electrode localization, ROI Grouping and Visualization

Electrode coordinates were obtained as previously described^23^. After transformation to MNI space, electrodes were visualized on the FSaverage surface. Group level results (**Figure. 2**) were obtained on an electrode-wise basis (using patient as repeated measure in **Figure. 2A**) given the near identical electrode configuration across patients. While electrodes were targeted to the same anatomic structures, there were differences in final affine-registered electrode positions due to residual anatomical variability and the need to adjust trajectories to avoid vasculature. The RMS variability in electrode coordinate locations is shown in supplemental **Figure S2**. ROIs were manually parceled and verified by two experts (RM, BS or RM, GB). ROIs of interest in the current study include 1) the ventromedial prefrontal cortex (vmPFC) across Brodmann areas 10, 24, 25, and part of 32, which contains the most anterior and ventral aspect of the anterior cingulate cortex (ACC); 2) the dorsal ACC (dACC) across area 24 and part of area 32; 3) the dorsal PFC (dPFC) across areas 8, 9, and 46; 4) the amygdala (AMY); and 5) the mid-temporal regions (MT, Temporal). Deterministic fiber tracking initiated from patient-defined bilateral vmPFC contacts highlights the engagement of three major affective network fiber systems, including the cingulum bundle (green), forceps minor (purple), and uncinate fasciculus (blue), in facilitating structural connectivity across ROIs (**Figure. S4**). The vmPFC region was transformed into ICBM152 space, and tractography was conducted in a population-averaged diffusion template available in DSI Studio (https://dsi-studio.labsolver.org). Individual fiber systems were isolated and identified through a clustering method in DSI Studio software, then visually-inspected.

## Results

### Aperiodic slope as an index for depression severity

We investigated whether variations in spectral slope could track depression severity across five patients undergoing simultaneous implantation of DBS and stereo-EEG electrodes for treatment of TRD as a function of an experimental clinical trial (NCT03437928). Stereo-EEG electrodes placed in target regions of the affective and reward processing networks (vmPFC, dPFC, dACC, AMY, MT; see methods for regional definitions) were used to record local field potentials (LFPs) during each completion of the CAT-DI^20^ across a 9-day in-patient hospital visit **(Table S1)**. Studies examining the aperiodic component typically constrain slope calculations to under ∼50 Hz^16,17^ to avoid the impact of line noise on the slope calculation, and given previous reports of large alpha and low beta features that give rise to the ‘knee frequency’^24,25^, we further imposed a limitation on the lower end of the range to help mitigate those known issues. Examination of the slope metrics within and across our patients confirmed greatest reliability and consistency in the 20-45 Hz sub-range. CAT-DI depression severity scores were then correlated with slope values at each electrode contact within patient **(Figure 1a)**, showing the strongest average correlations within the vmPFC. Each patient showed a flattening (reduction of magnitude of negative value) of the aperiodic slope in the ‘biomarker’ region (vmPFC) between the highest [70-96] and lowest [27-68] depression severity scores from the entire in-patient stay **(Figure 1b)**. Aperiodic slope was averaged across all electrodes within the vmPFC region and related to depression severity scores in each patient in **Figure 1c**, yielding r=[-0.58 to −0.34] (p=[0.0004 to 0.017]), indicating significantly flatter slope during more euthymic (less depressed) mood states in every subject. We also examined the relationship of aperiodic slope to depression severity when averaging across all electrodes and for individual ROIs (see ROI definition in **Figure 2a** and supplemental results in **Figure S1**). Control analysis using permutation testing confirm these findings, and a binomial cumulative distribution function shows a significant slope-severity relationship within each patient (p=0.00 to 7.51e-11; **Table S2**).

**Figure 1.**
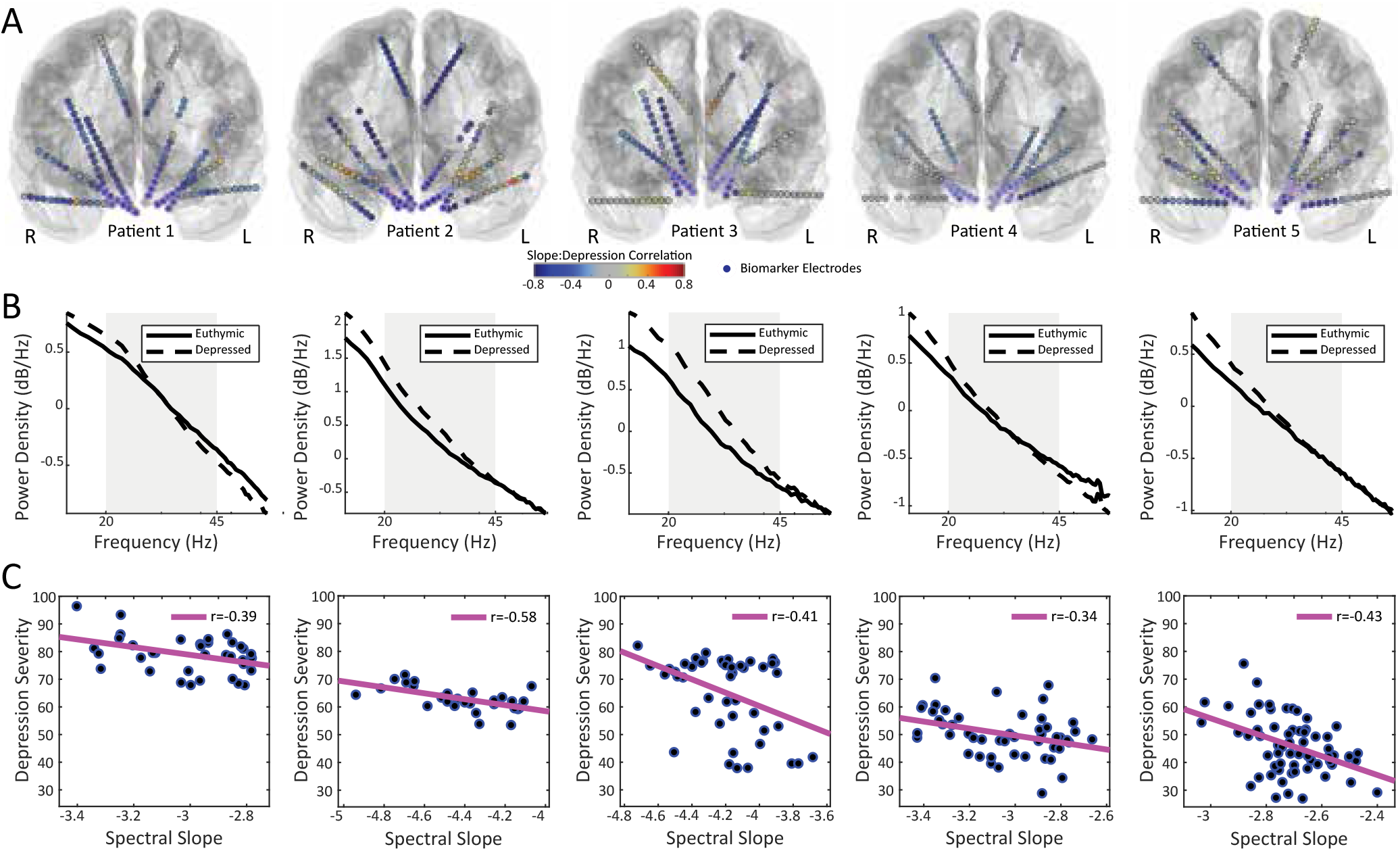
Aperiodic slope as depression severity biomarker. **(A)** The aperiodic slope was computed for each depression severity (CAT-DI) assessment block per electrode and correlated with the depression severity scores. Electrode coloring indicates slope-depression correlation magnitude; magenta outlines designate electrodes belonging to the putative ‘biomarker’ ventromedial PFC region of interest (vmPFC ROI). **(B)** Power spectra averaged across the ‘biomarker’ vmPFC region plotted for the highest (most depressed) and lowest (most euthymic) CAT-DI scores for each patient, showing that as slope becomes less negative (flatter) depression symptoms seem to alleviate. **(C)** Depression severity vs slope (averaged over vmPFC) for all scores collected during the 9-day in-patient monitoring period.

### Affective-regional specificity of the aperiodic biomarker across patients

Typically, locations for sEEG electrode placement are idiosyncratic to clinical hypotheses for individual patients undergoing invasive monitoring for epilepsy, resulting in a high degree of spatial variability in sampling. Here, sEEG target regions were determined *a priori* and trajectories are only altered slightly based on anatomical variation and vasculature, thus allowing for consistent sampling across regions. This offers a unique opportunity for a group-level electrode co-registration, such that each electrode site can be averaged in space across patients **(Figure 2a)**. Despite some variability in location after co-registration (**Figure S2**), combining correlations across electrodes again indicates that the largest effect sizes for the slope-depression correlations were consistently located in the vmPFC (**Figure 2b**). To further characterize any regional specificity, electrodes were grouped by subregion (vmPFC, dPFC, AMY, dACC, and MT). A violin plot showing point aggregation represents the spread of correlations for all electrodes across all subjects grouped by ROI, and indicates the aperiodic biomarker for mood is strongest in the vmPFC (slope vs. depression severity correlation median=-0.35, IQR=0.18) and weakest in lateral temporal contacts (median=-0.06, IQR=0.24) **(Figure 2c)**. A one-way ANOVA revealed a significant effect of group on the response variable, F(4,540)=16.03, p<3e-12. Notably, while the overall effect in the amygdala showed a moderate relationship between aperiodic slope and depression severity, group-wise visual inspection revealed a consistent lateralization in the amygdala; the left amygdala showed a strong, negative relationship between aperiodic slope and depression severity over subjects (r=-0.35±0.19, p=4e-6), whereas the right amygdala showed little to no effect (r=-0.17±0.29, p=0.02), as demonstrated in **Figure 2b**. To confirm that our *a priori* frequency range was appropriate for slope calculations, we next correlated depression severity scores with vmPFC aperiodic slope across a broad frequency range (f_low_=1 to 99 Hz, f_high_= f_low_ to 100 Hz). The *a priori* range spanned the beta and low gamma ranges (white square, 20-45 Hz; **Figure 2d**) was well within the broad range of frequencies showing a significant relationship between slope and depression severity. The boundary designated by the darkest blue region in **Figure 2d** indicates there is a set of frequency ranges which may be more optimal than (yet still similar to) the *a priori* range for detecting the slope-severity relationship.

**Figure 2.**
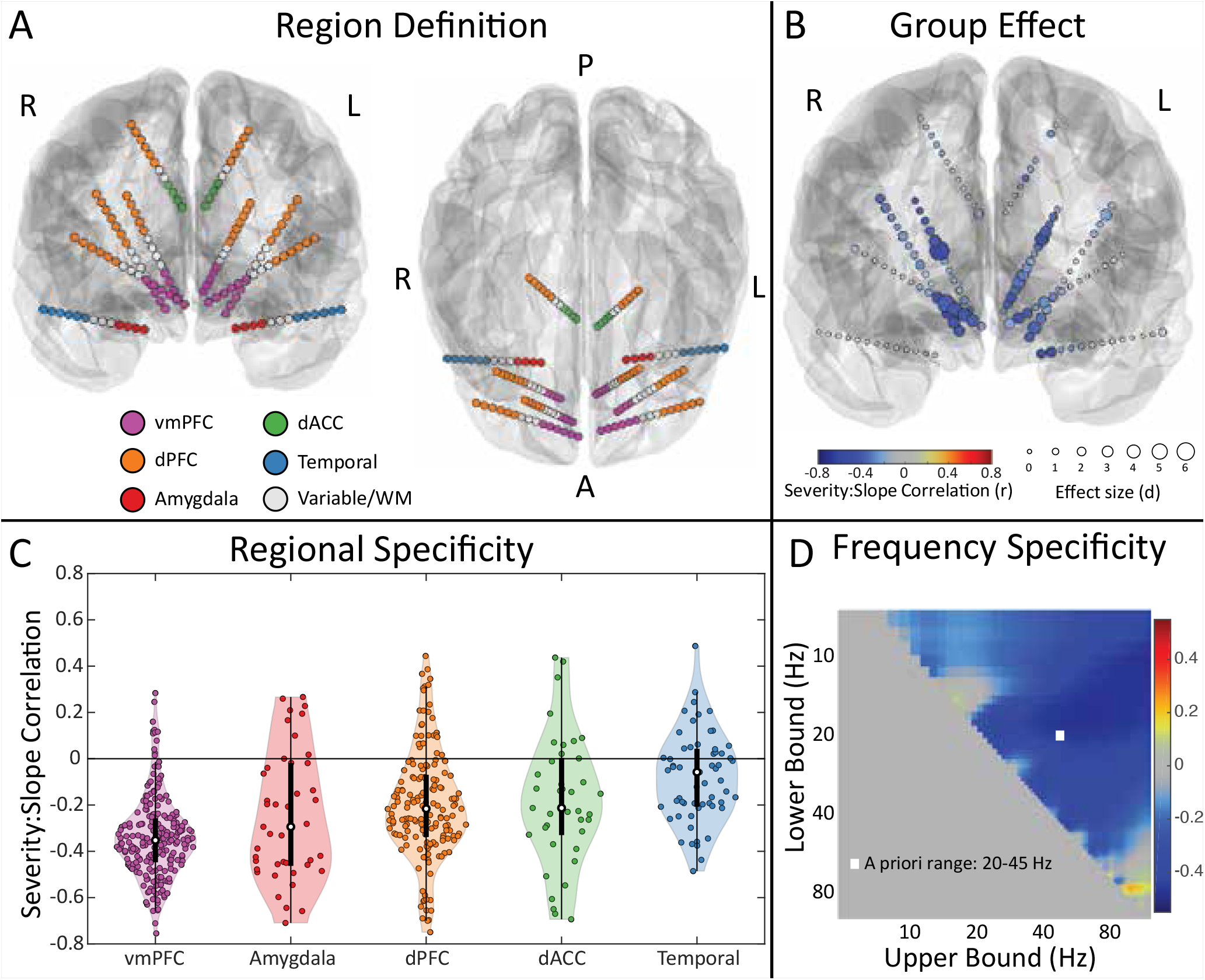
Biomarker regional and frequency specificity. **(A)** Electrodes were divided into five ROI groups indicated by fill color. **(B)** Similar electrode placements allowed inter-subject electrode registration. Electrode fill color shows the average correlational value across subjects (r = −0.5 to 0.06), and electrode size indicates the effect size of the relationship at each contact (Cohen’s d range = −6.5 to 0.3). **(C)** The distribution of regional depression severity:slope correlations indicates that the vmPFC exhibits the strongest relationship with depression severity score (median = −0.35, IQR = 0.18), and that the temporal contacts exhibit the weakest relationship (median r = −0.06, IQR = 0.24). Each marker represents one electrode from one subject. **(D)** Severity-slope correlations were calculated for every frequency range from 1 to 100 Hz and averaged in the vmPFC region. The white square designates the location of the a priori range (20-45 Hz).

**Figure 3.**
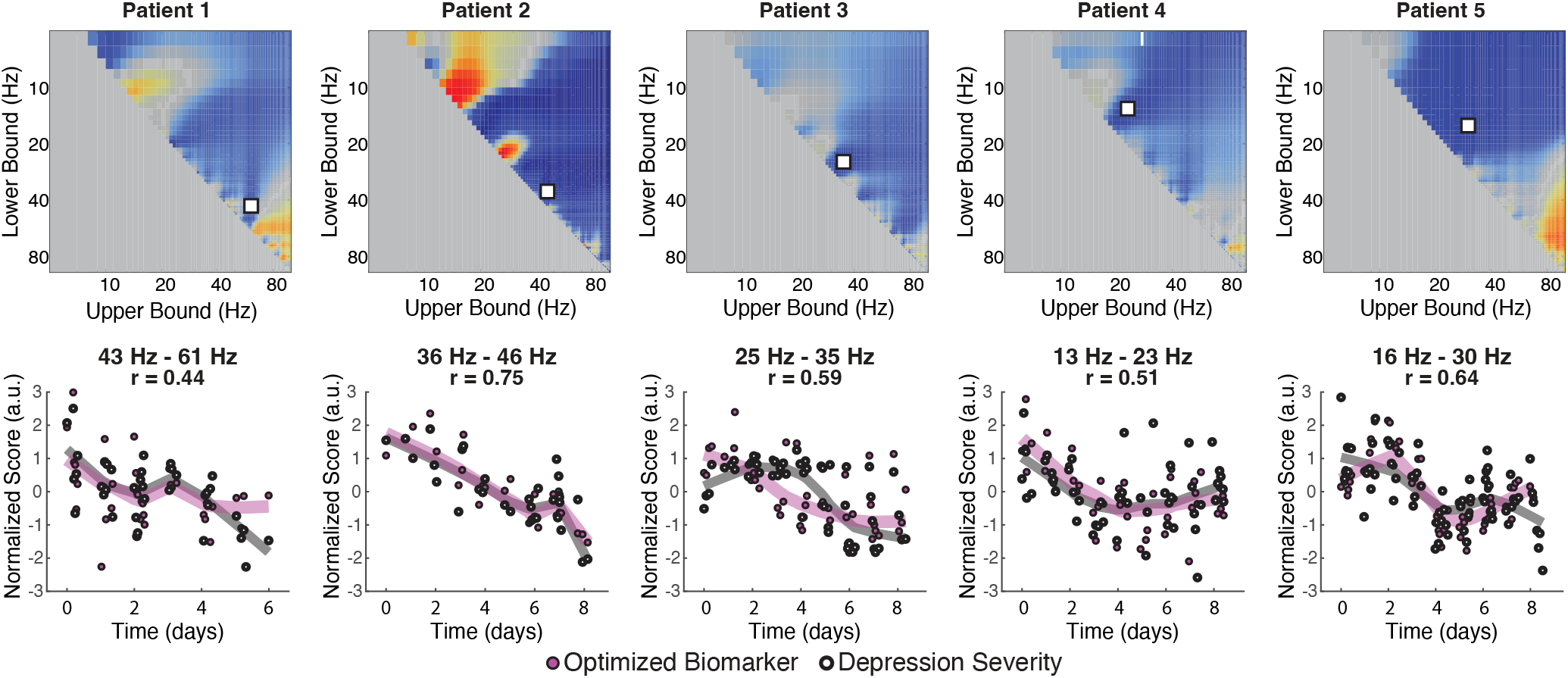
Optimized depression severity tracking. **(A)** Aperiodic slope was calculated for each possible frequency range (1-100 Hz) within the vmPFC for each patient and correlated with depression severity scores. The frequency range for a given correlation is identified by the intersection of the lower (y-axis) and upper (x-axis) bounds. **(B)** The optimal frequency range indicated by a white square in **(A)** was used to calculate an optimized aperiodic slope estimate (magenta points) for each depression severity assessment (white points). Both sets of scores were individually Z-scored then plotted as a function of time. Magenta and black lines show robust LOWESS moving average estimates (span = 0.5).

### Patient-specific biomarker optimization

We next sought to examine the strength with which an optimized biomarker would track each individual’s mood over the in-patient stay. We examined the relationship between each vmPFC spectral slope and depression severity for each combination of lower and upper frequency bound (similar to **Figure 2c**) for each patient (**Figure 3a**). The optimal spectral slope computation was then illustrated as a function of time along with depression severity in **Figure 3b** yielding an improved depression severity biomarker (r=0.44 to 0.75, p-value range=0.006–1.8e-6).

### Biomarker free of diurnal fluctuations

Given previous findings suggesting that aperiodic slope may be a proxy for arousal state^26^, we aimed to identify if the fluctuations seen in the slope biomarker were due to circadian changes in arousal throughout the day. We binned severity and slope by time of day to inspect the relationship between depression severity and spectral slope (both normalized by Z-scoring within day, **Figure S3**) and did not find any significant diurnal relationship for either variable.

## Discussion

The poor treatment success rate in MDD^1^ emphasizes the necessity for individualized therapeutic approaches. A task-independent physiological biomarker of mood would facilitate this shift in three ways by: 1) enabling faster tracking of mood changes compared to the typical 2-week assessment period^27,28^, 2) passively tracking mood to mitigate patient biases arising from social, physical, and emotional factors^3,29–31^, and 3) serving as a comprehensive metric that remains consistent across frequency range, brain region, and patients. In this study, we demonstrate that the aperiodic component of the power spectrum effectively tracks mood changes over minutes to hours, displaying both global and flexible regional and frequency specificity across patients. Moreover, the aperiodic slope is easily calculated, and the relationship between slope and depression severity is visually evident in each patient, obviating the need for machine learning techniques. To our knowledge, this study is the first to identify a robust biomarker that fulfills all three criteria.

### Periodic and Aperiodic Biomarkers

Previous studies searching for a physiological biomarker have primarily focused on periodic, oscillatory data, eliminating the aperiodic component by normalizing power band signals to a baseline period for trial-level analyses in task-based designs^32^, or by regressing out the negative linear slope with techniques such as IRASA^33^ and FOOOF^34^. These studies have identified potential biomarkers for treatment response and mood in various frequency bands (theta^11^, alpha^9,10^, beta^7,9^, high and low gamma^7,9,35^) across affective regions. However, the wide variety of reported frequency bands may all be driven by a common underlying phenomenon – a change in the power law slope across all frequency bands. Previous studies have necessarily relied on group averages (i.e., depressed vs. non-depressed) and often require advanced statistical modeling for detection and prediction. The variability in anatomical location and frequency range of oscillatory biomarkers limits their clinical feasibility for closed-loop control in treatments like deep brain stimulation^10^. Our group adopted a canonical frequency band-based approach and utilized linear state-space modeling, revealing a consistent ‘high-high low-low’ pattern in prefrontal regions across an initial study cohort of 3 participants^7^ (patients 1-3 in the current study). This pattern indicated that increased high-frequency band power coupled with reduced low-frequency band power in the ACC correlated with decreased depression severity. However, this approach neglected the removal of the aperiodic slope, potentially suggesting that the observed pattern reflects shifts in the aperiodic component rather than the oscillatory components themselves^34^. The specific oscillatory bands identified as predictors of depression severity varied between subjects^7^, further highlighting the inconsistency of using oscillatory signals as biomarkers.

More recently, Alagapan et al. (2023) leveraged a neural network classifier in a data pool of five TRD patients to identify discriminative frequency band components and capture differences between the TRD ‘sick’ and ‘stable’ response states^9^. Though a powerful approach, this study showed that their spectral discriminative component (SDC) was associated with changes in many spectral frequency bands simultaneously within the subcallosal cingulate cortex (SCC). Interestingly, this change typically showed an increase in high frequency and a decrease in low frequency spectral components (‘high high’, ‘low low’), with some variability in specific feature fluctuations between hemispheres. This finding is in line with prior reports from in the ACC of TRD patients^7^, and suggests that broad, simultaneous changes in spectral components may actually reflect a shift in the aperiodic component of the PSD. Excitingly, since the aperiodic slope can capture the overall balance of activity across frequency bands, it is inherently achieving the reduction in dimensionality that the SDC is meant to represent, without requiring weeks-to-months of data collection per patient. Furthermore, the use of aperiodic slope as in the present study requires only a few minutes of recording, as opposed to the use of other candidate features^9^ which are only able to predict depression severity when trained on many weeks to months of data, thus limiting their utility for tracking depression in sparser or more immediate sampling scenarios.

Assessing the aperiodic component overcomes limitations observed in previously identified biomarkers. The relationship between slope and depression severity is evident even when averaging across all electrodes, and the strongest correlations with the biomarker are consistently observed in the vmPFC across patients. While the optimal frequency range varies per individual at the group level, a broad range of frequency pairs (darkest blue area in **Figure 2d**) produces a reliable feature for detecting the slope-severity relationship. Furthermore, the calculation of the aperiodic slope is straightforward, involving a simple three-step process (decompose, log-transform, linear-fit), and the current study has demonstrated a robust relationship from only 2-4 minute-long recordings. These parameters allow for real-time assessment during recording and stimulation paradigms. Future studies can integrate the aperiodic metric as a measure of moment-to-moment mood changes, incorporating specific region-of-interest (ROI) and electrophysiological parameters from the outset, while retaining flexibility to modify frequency and regional parameters for each individual as more data are accumulated.

Prior studies and the current study suggest the possibility of leveraging differences in oscillatory activity between patients to adapt diagnoses and treatments to each patient^7,9^. While the aperiodic component provides consistency for tracking depression severity over time and across patients, specific oscillatory bands may be valuable for decoding depression subtypes and capturing distinct behavioral phenomena. Recent language studies highlight the importance of both aperiodic and oscillatory components in cognition, with slope tracking higher-order cognitive processing and oscillatory profiles reflecting sub-domains of language^36^. Similarly, aperiodic slope in affective hubs may represent an ‘affective gain’ and capture the overall affective network tone of an individual, while distinct oscillatory patterns manifest as electrophysiological markers of mood symptom sub-domains (i.e. reward or affective processing dysfunction, hypersomnia or insomnia, alexithymia, etc.). Evidence suggests that oscillatory balance seen in the aperiodic component could reflect overall affective processing rather than specific sub-domains^37^.

An additional advantage of the current finding is that it suggests a new axis by which to conceptualize the pathophysiological state underlying depression. By thinking of depression as an imbalance of excitation and inhibition, in neural circuits essential for maintaining mood, future research efforts may aim at optimizing this balance, whether by pharmaceutical, electroceutical or other means. Indeed, excitatory/inhibitory (im)balance has long been implicated in neural pathology, including in epilepsies, schizophrenia, autism spectrum disorders, and other neuropsychiatric conditions^38,36^.

### Regional and Frequency Specificity of the Aperiodic Biomarker

Prior to this clinical trial, the search for biomarkers of depression focused on non-invasive methods^39^ and recordings from sub-optimal anatomical sites (white matter) and in the context of ongoing stimulation using complex artifact rejection techniques such as common mode rejection^40^. The current clinical trial captured recordings from grey matter through sEEG electrodes placed in tandem with DBS sensing and stimulating electrodes, enabling enhanced sensitivity to local field potential (LFP) activity, which has perhaps provided the analytical scope for the current discovery. Grouping sEEG electrodes into five ROIs, we observed a consistent phenomenon across a broad frequency range and most anatomical regions that robustly tracked mood (**Figure 2, S1**). While each patient and region show slight variation in the optimal frequency range for calculating slope (**Figure 2c, 3a, 3S**), the *a priori* (20-45 Hz) frequency range was robust across patients and regions. This ‘global’ characteristic of the aperiodic slope gives the option to track depression severity by averaging aperiodic slope over all electrodes within this arbitrarily defined wide frequency range before individualized parameters are acquired (**Figure S1**).

Despite the aperiodic biomarker’s global nature compared to oscillatory biomarkers (**Figure 1, S1**), the strongest relationship between depression severity and aperiodic slope was consistently observed in the ventromedial prefrontal cortex (vmPFC). This aligns with existing literature highlighting the importance of the ventromedial prefrontal region in reward, social, and affective processing^41^, as well as its position as a central hub in the default mode network^42^ with connections to hubs of the affective and salience networks^43^. Notably, the most anterior and ventral portion of the ACC (subgenual/subcallosal ACC) which was grouped in this study’s vmPFC ROI, is a well-known target of DBS therapies for depression^42^ because of its ideal location along the prominent white matter tract (the cingulum bundle) responsible for innervating a vast portion of cortical and subcortical regions involved in mood modulation. We visualized the recruitment of affective network-related white matter tracts through whole brain deterministic tractography initiated from the vmPFC region, as defined in this study (**Figure S4**). Contacts located within the vmPFC recording sites exhibited dense innervation of the cingulum bundle, forceps minor, and uncinate fasciculus. These fiber systems represent three of the four white matter bundles targeted in neurosurgical planning for subcallosal cingulate deep brain stimulation for TRD^43^. Further, the cingulum bundle and forceps minor have been identified as the likely stimulation pathways producing stable, efficacious responses in SCC DBS^44^. Together, the cingulum bundle, forceps minor, and uncinate fasciculus facilitate structural connectivity from the vmPFC to ACC, DPFC, and amygdala regions^45,46^. Thus, the “biomarker region” in the current study may represent a hub region of integration across multiple streams of input and output of the affective networks (**Figure S4**).

Interestingly, the dorsal portion of the anterior cingulate cortex is also an important hub of affective processing, with structural and functional connections with the subgenual cingulate cortex (SCC) and ventral capsule/ventral striatum (VC/VS) targets in DBS therapies^43^, and has been previously highlighted as a potentially sensitive region in our cohort^7^. While it is surprising that this region did not show a stronger relationship with depression severity, we did observe the contacts in the dACC had the greatest variability in trajectory and placement across our patients (**Figure S2.**). Future studies should reexamine the aperiodic signature in the dACC using a greater sampling along the dACC structure to help characterize this complexity.

A strong relationship between depression severity and aperiodic slope was observed in the left, but not the right amygdala. Asymmetry of amygdala dysfunction is commonly reported in MDD, though the specific mechanisms remain unclear^47,48^. One longitudinal study revealed an emotional processing bias which was normalized after effective treatment in people with depression, which was specifically significant in the left amygdala^48^. Additionally, patients with depression demonstrated a significant negative correlation between activity in the left amygdala during positive memory recall and HAM-D score^49^. Considering our findings, it is possible that the left amygdala could exhibit greater sensitivity to depressive symptomatology than the right.

### Limitations and Future Directions

The aperiodic slope meets and exceeds three important criteria essential to an appropriate biomarker in that it can track depression severity on the order of minutes to hours, be passively collected during patient monitoring, and is highly robust in frequency and anatomical region across patients. However, the current study was limited to the in-patient phase of the clinical trial. For this reason, we were not able to identify whether the aperiodic slope, which was collected intracranially in this study, is generalizable to non-invasive measures. Future studies must investigate whether non-invasive measures (scalp EEG, fMRI), or peripheral nervous system markers (EKG, skin conductance), carry the same power law biomarker relationship across depression severity^32^.

Furthermore, given that the aperiodic slope is suggested to act as an index for arousal^26^, it is difficult to disentangle if this metric is a true biomarker for mood, which is associated with changes in arousal, or if it is a biomarker for arousal, which is associated with mood. The latter implies that in states of high or low arousal which are independent of trait-mood, during different wake or sleep states for example, aperiodic slope would not maintain its validity as a biomarker. Our results showing the daily progression of mood and scores across the course of a day suggests that the relationship we see here is not diurnally driven (**Figure S3**), thus providing evidence that the biomarker is tracking mood, or at least, arousal as it relates to mood and not as it relates to the circadian cycle. Previous studies have shown that aperiodic slope during sleep can distinguish depressed from non-depressed populations^16^, though future studies are needed to examine the differences in aperiodic slope between sleep and wake.

Future studies must investigate whether this biomarker effectively tracks changes in mood on the order of minutes to seconds, as it does with auditory stimuli. This is likely the case, as aperiodic slope shifts on the order of milliseconds to seconds following the presentation of salient auditory stimuli^18^; thus, future studies should address if aperiodic slope in the affective network can be used as an instantaneous measure of mood. Further studies must also examine how this relationship exists in responders vs. non-responders to treatment, if it can track other arousal-mood pathologies including anxiety disorders and PTSD, and how this relationship might shift across age, which is already known to affect the aperiodic slope component of the PSD^13^.

## Conclusion

This study presents compelling evidence for the aperiodic slope as a reliable and temporally sensitive physiological biomarker for MDD, and especially in treatment-resistant depression (TRD). Our results corroborate previous findings about the aperiodic slope’s responsiveness to long-term mood changes following MDD treatment and further demonstrate its efficacy in tracking mood changes at high temporal resolution compared with conventional assessments. In contrast to the inconsistent oscillatory markers, the aperiodic slope provides a consistent, task-independent, and visually discernible measure of depression severity across patients, thereby eliminating the need for complex machine learning techniques. This consistent and highly temporally-resolved biomarker has the potential to revolutionize MDD treatment by facilitating efficient screening of therapeutic approaches, individualizing treatment strategies, and enabling active control of the dysfunctional affective system in TRD. Future research can further investigate the potential of integrating the aperiodic metric as a real-time measure of mood changes and explore the co-contributions of the aperiodic and oscillatory components to the understanding of depression and its subtypes. The aperiodic slope can potentially open new avenues in the search for a reliable biomarker for psychiatric disorders, contributing to the move towards precision psychiatry^50^.

## Supporting information

Supplement_FigureLegends

## Data Availability

All data, code, and materials used in the analysis are available to any interested researcher for the purpose of reproducing or extending the analysis. Primary data are also hosted on the National Institutes of Mental Health Data Archive (NDA), and analysis data frames are posted on the Data Archive of the BRAIN Initiative (DABI).

## Acknowledgements

The authors would like to thank the patients and their families for their participation in the research. We would like to thank Cameron McIntyre and Angela Noecker for their support in neuroimaging visualization for figure S4 and cover art submission. We would like to thank the intracranial monitoring unit staff (Luis Escudero, Katrina Reichardt), and collaborators at Boston Scientific (Allen Stutes, Steven Carcieri, Mahsa Malekmohammadi, Richard Mustakos), as well as the following individuals for their support of the project: Ricardo Najera, Adrish Anand, Ron Gadot, Ethan Devara, Huy Dang, Nabeel Diab, Sameer Rajesh, Katherine Kabotyanski, K., Robert Petrovic, Anthony Allam, Sandresh Reddy, and Gabriel Reyes. We would like to thank Robert Gibbons and Yehuda Cohen for their support of the acute application of the CAT-DI tool.

## Author contributions

Conceptualization: KB, CH, MM

Methodology: CH, KB, JA, AW, MM

Investigation: MM, JX, EB, AW, JA

Visualization: CH, ID, AM, MM, BP, RM, AN, CM, GB, BS

Funding acquisition: KB, SS, NP, DB, JC, WG

Project administration: VP

Supervision: KB, SS

Writing – original draft: MM, CH, KB, XF, ID

Writing – review & editing: MM, CH, KB, NP, DAB

