## Supplement_FigureLegends for "Aperiodic neural activity is a biomarker for depression severity"

Affiliations:

Baylor College of Medicine

S101D Smith Biomedical Research Building

One Baylor Plaza

Houston, TX 77030

One sentence summary: The aperiodic component of the power spectral density robustly tracks depression severity on the order of minutes to hours.

Supplementary Materials

Figure Legends

Figure. S1. Depression severity vs slope correlation across five regions of interest.

Figure. S2. Electrode placement variability across subjects.

Figure. S3. Non-diurnal slope-severity relationship.

Figure. S4. Connectivity of biomarker region to affective network structures.

**Supplementary Figure 1.** Depression severity vs slope averages for all scores collected during the 10-day in-patient monitoring period. The first row shows comparisons of depression severity to aperiodic slope averaged over all electrodes. Subsequent rows compare to averages across each ROI subregion (see main text Figure 2 for definitions).

**Supplementary Figure 2. Similarity of electrode placement across subjects.** Marker size indicates the root-mean square dispersion of each electrode across patients after registration across similar trajectories. Note reference/ground electrodes (left dorsal region) were omitted from all analyses leading to focally decreased dispersion statistics for some contacts.

**Supplementary Figure 3. Relationship of spectral slope and depression severity to time of day.** Depression severity and spectral slope (averaged over vmPFC region) were sorted by day (dividing days at 0600) and Z-scored within day. Neither measure showed a significant relationship across patients to time of day.

**Supplementary Figure 4. Connectivity of biomarker region with affective network structures.** Whole brain deterministic tractography was initiated from the vmPFC biomarker region to estimate the structural connectivity of this region to other limbic structures. **(A)** Regions of interest (ROIs) used in the current study observed in one cerebral hemisphere: biomarker ventromedial prefrontal (vmPFC) region (magenta), dorsal prefrontal (dPFC) (orange), anterior cingulate (ACC) (green), amygdala (red), and mid-temporal (blue). **(B)** ROIs overlaid on vmPFC-affective network connectivity in the left hemisphere. **(C)** Bilateral affective network connectivity demonstrating innervation patterns from the uncinate fasciculus (blue), forceps minor (purple), and cingulum bundle (green), isolated through tract clustering algorithm in DSI Studio. Recruitment of these three major fiber systems contributes substantially to treatment efficacy in deep brain stimulation, and appear to work by unifying affective function across the distributed network space of the dorsal ACC, dPFC, amygdala, and vmPFC.
