## Supplementary figures and images for "Aperiodic neural activity is a biomarker for depression severity"

### Supplemental Figure 1

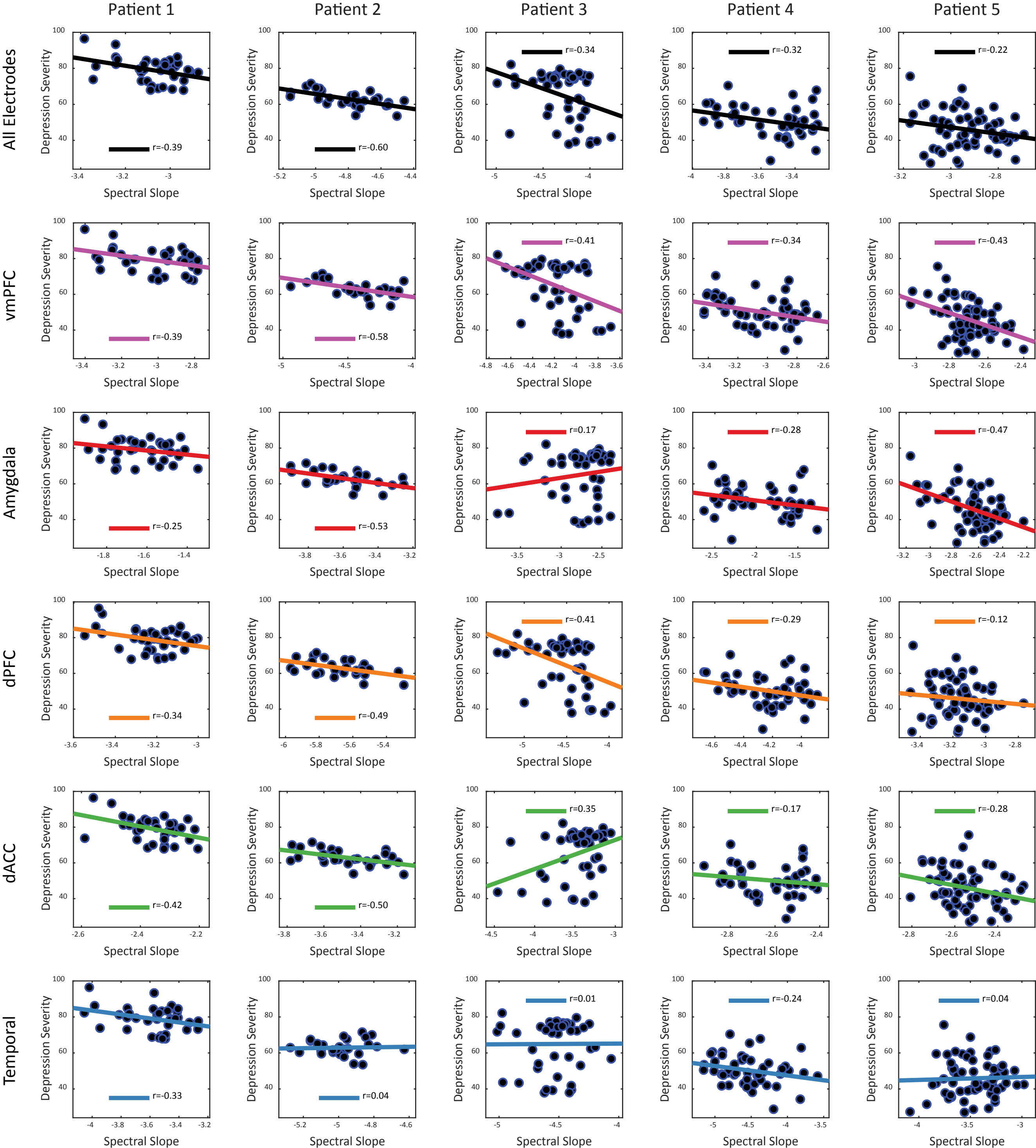

### Supplemental Figure 2

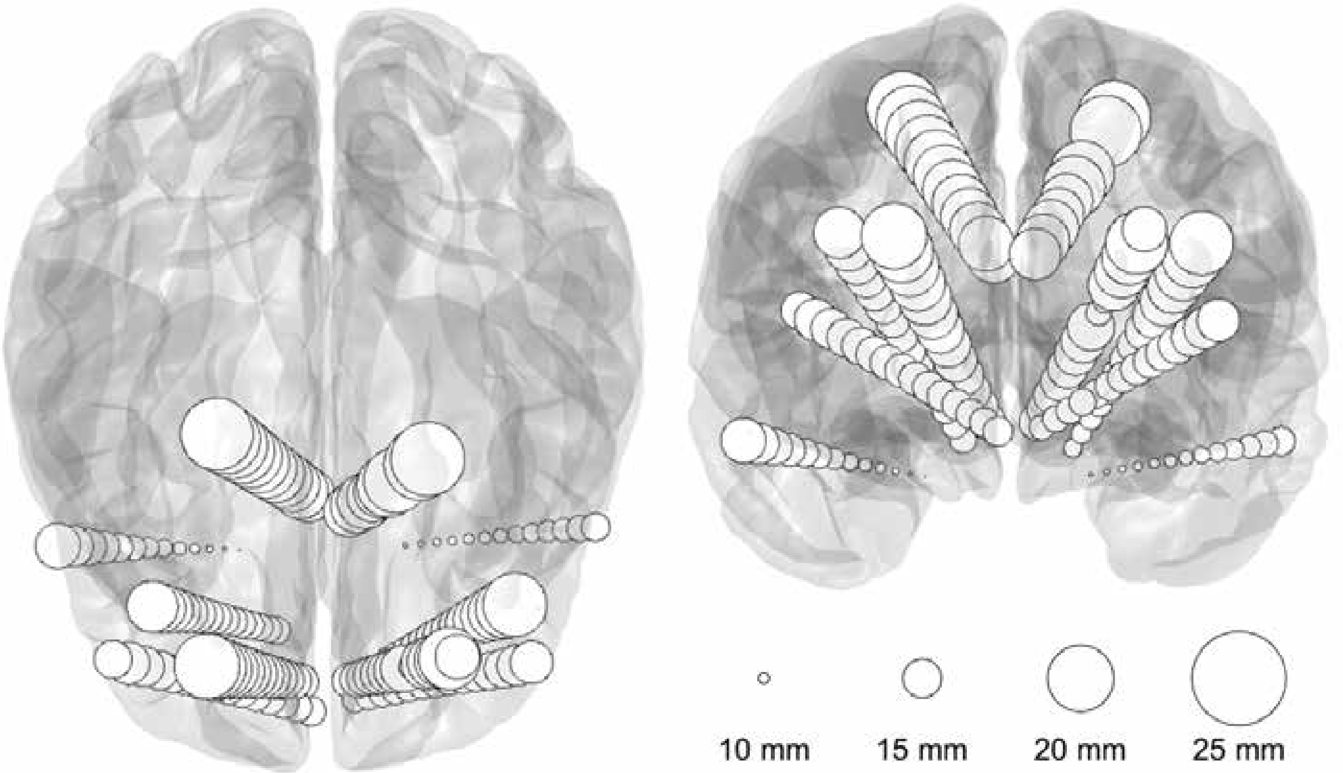

### Supplemental Figure 3

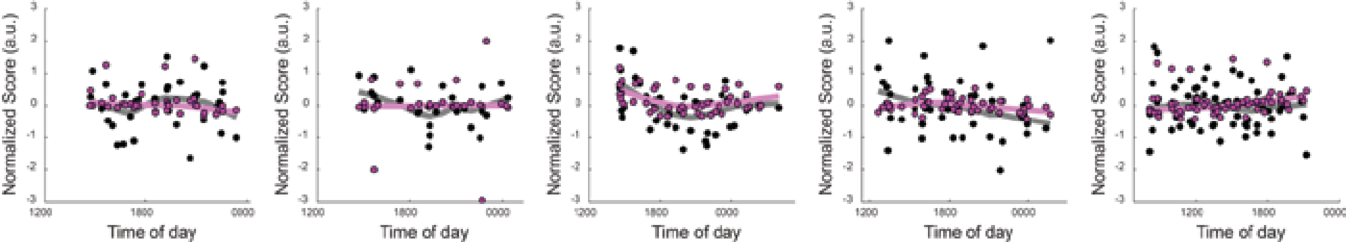

### Supplemental Figure 4

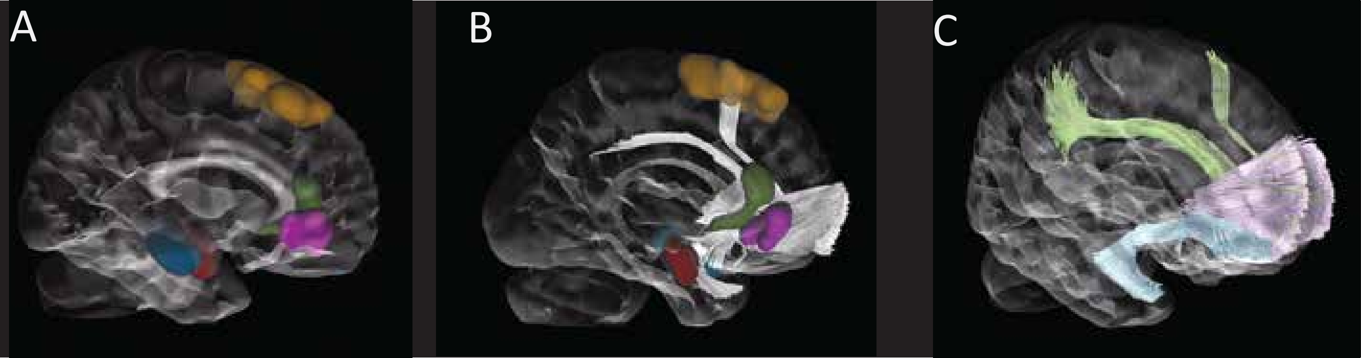
