## Supplemental Table 1 for "Aperiodic neural activity is a biomarker for depression severity"

**Table S1.** Frequency and Average length of CAT-DI assessments per patient.

| | Patient 1 | Patient 2 | Patient 3 | Patient 4 | Patient 5 |
|---|---|---|---|---|---|
| Assessment Count | 37 | 30 | 48 | 50 | 65 |
| Assessment Length [sec] Mean(SD) | 201.80(84.38) | 75.77(32.38) | 120.30(80.80) | 101.96(31.96) | 114.27(48.75) |
