## Supplemental Table 2 for "Aperiodic neural activity is a biomarker for depression severity"

**Table S2.** Permutation testing for slope-severity correlation per channel.

| | Patient 1 | Patient2 | Patient 3 | Patient 4 | Patient 5 |
|---|---|---|---|---|---|
| Total Channels | 179 | 176 | 184 | 180 | 172 |
| Original Significant Channels | 87 | 70 | 68 | 29 | 62 |
| Permutation Test Significant Channels (Positive Correlation/ Negative Correlation) | 87(1/86) | 69(3/66) | 67(7/60) | 25(0/25) | 63(24/39) |
| Patient Significance (Binomial Result p-value) | 0.00 | 0.00 | 0.00 | 1.26e-06 | 0.00 |
